# Characterization of Per- and Polyfluoroalkyl Substance (PFAS) concentrations in a community-based sample of infants from Samoa

**DOI:** 10.1101/2023.11.10.23298357

**Authors:** Lacey W. Heinsberg, Shan Niu, Kendall J. Arslanian, Ruiwen Chen, Megha Bedi, Folla Unasa-Apelu, Ulai T. Fidow, Christina Soti-Ulberg, Yvette P. Conley, Daniel E. Weeks, Carla A. Ng, Nicola L. Hawley

**Affiliations:** Department of Human Genetics, School of Public Health, University of Pittsburgh, Pittsburgh, Pennsylvania, USA; Department of Civil and Environmental Engineering, Swanson School of Engineering, University of Pittsburgh, Pittsburgh, Pennsylvania, USA; Department of Social and Behavioral Sciences, Yale University School of Public Health, New Haven, Connecticut, USA; Obesity, Lifestyle and Genetic Adaptations Study Group, Apia, Samoa; Samoa Ministry of Health, Apia, Samoa; Department of Health Promotion and Development, School of Nursing, University of Pittsburgh, Pittsburgh, Pennsylvania, USA; Department of Biostatistics, School of Public Health, University of Pittsburgh, Pittsburgh, Pennsylvania, USA; Department of Chronic Disease Epidemiology, Yale University School of Public Health, New Haven, Connecticut, USA

**Keywords:** Pacific Islands, Pacific Islander, Environmental health, Exposure science, Cord blood, Dried blood spots, Infant exposure

## Abstract

Per- and polyfluoroalkyl substances (PFAS) are persistent contaminants with documented harmful health effects. Despite increasing research, little attention has been given to studying PFAS contamination in low- and middle-income countries, including Samoa, where there is more recent modernization and potential window to examine earlier stages of PFAS exposure and consequences. Using data and biosamples collected through the *Foafoaga o le Ola* (“Beginning of Life”) Study, which recruited a sample of mothers and infants from Samoa, we conducted an exploratory study to describe concentrations of 40 PFAS analytes in infant cord blood collected at birth (n=66) and dried blood spots (DBS) collected at 4 months post-birth (n=50). Of the 40 PFAS analytes tested, 19 were detected in cord blood, with 11 detected in >10% of samples (PFBA, PFPeA, PFHpA, PFOA, PFNA, PFDA, PFUnA, PFTrDA, PFHxS, PFOS, and 9Cl-PF3ONS); 12 analytes were detected in DBS, with 3 detected in >10% of samples (PFBA, PFHxS, and PFOS). PFAS concentrations were generally lower than those reported in existing literature, with the exception of PFHxS, which was detected at higher concentrations. In cord blood, we noted associations between higher PFHxS and male sex, higher PFPeA and residence in Northwest ‘Upolu (NWU) compared to the Apia Urban Area (AUA), and lower PFUnA and 9Cl-PF3ONS with greater socioeconomic resources. In DBS, we found associations between higher PFBA and greater socioeconomic resources, and between lower PFBA and PFHxS and residence in NWU versus AUA. However, the latter association did not hold when controlling for socioeconomic resources. Finally, we observed associations between nutrition source at 4 months and DBS PFBA and PFHxS, with formula- or mixed-fed infants having higher concentrations compared to exclusively breastfed infants. This study presents the first evidence of PFAS contamination in Samoa. Additional work in larger samples is needed to identify potentially modifiable determinants of PFAS concentrations, information that is critical for informing environmental and health policy measures.

## 1. INTRODUCTION

Per- and polyfluoroalkyl substances (PFAS) are a group of man-made chemicals that emerged in the 1940s. Lauded for their unique properties of resistance to oil, grease, water, and heat, PFAS became ideal candidates for a wide range of applications, such as stain- or water-resistant fabrics, carpeting, fast food packaging, non-stick cookware, and fire-fighting foams.^1, 2^ While the chemical structures of PFAS are responsible for these desirable properties, they unfortunately also enable many PFAS to mimic fatty acids.^3, 4^ As a result, PFAS are bioavailable and many bioaccumulate and interfere with normal physiological processes.^5^

In fact, alarming evidence has emerged linking PFAS exposure to a myriad of negative health effects across the lifespan, including increased cholesterol levels, liver dysfunction, lower birth weight, elevated blood pressure, endocrine disruption, immune hazards, and more.^6, 7^ PFAS contamination is observed even in remote locations. For example, in the Faroe Islands, human biomonitoring studies have shown the presence of PFAS in blood and associated adverse health effects, particularly immune dysfunction.^8^ Some of the most common sources of human exposure include proximity to airports or military bases (where PFAS-containing fire-fighting foams have been historically used), consumption of PFAS-contaminated water or food (e.g., fish), and use of products made from PFAS.^9^

Despite the growing body of research on PFAS, the majority of work to date has predominantly focused on characterizing PFAS levels and health effects in higher-income countries. Even in those regions, the failure of policies to keep up with the rapid pace of scientific findings has led to concerning instances of ‘regrettable substitution’—the replacement of banned PFAS chemicals with structurally similar alternatives that may be equally harmful, or potentially even worse.^10^ This practice makes it exceedingly challenging to effectively monitor and manage PFAS and predict their public health consequences accurately.^11^

Unfortunately, research efforts on PFAS in low- and middle-income countries, including Samoa, have been limited^12–15^ despite the rapid modernization and increasing prevalence of chronic disease in these regions.^16, 17^ Unlike higher-income countries where PFAS contamination has been a known issue for decades, Samoa’s more recent development offers a unique opportunity to examine the earlier stages of exposure and its consequences. This temporal advantage could enable us not only to understand the evolution of PFAS-related health effects over time, which is crucial for informed policymaking and public health interventions, but also to begin to address the global issue of environmental justice across historically excluded communities.

Therefore, the purpose of this exploratory study was to begin to address this critical knowledge gap by characterizing PFAS concentrations in cord blood, collected at birth, and dried blood spots (DBS), collected at 4 months post-birth, in infants from Samoa, and examine associations with participant factors such as sex, geographic region of residence, socioeconomic resources, and nutrition source. The focus on infants is particularly significant, as early-life exposure to PFAS may have far-reaching consequences on health and development into adulthood.^18^ In light of the potential risks associated with PFAS exposure, understanding the levels of these substances around the world is of paramount importance to support continued research to improve health equity and to inform policy measures to safeguard public health globally.

## 2. MATERIALS AND METHODS

### 2.1 Study overview, design, and setting

Data were derived from a longitudinal observational study of mother-infant dyads from Samoa. Participants were recruited through the 2017-2019 *Foafoaga o le Ola* (“Beginning of Life”) parent study, which focused on identifying factors in pregnancy and the early postpartum period that contributed to non-communicable diseases in the Samoan setting.^19^ Dyads (n=161) were enrolled before birth at the antenatal care clinic of the Tupua Tamasese Meaole (TTM) Hospital in Apia, Samoa. At only approximately 2,800 km^2^, Samoa is home to approximately 222,000 people.^20^ Samoa is geographically isolated and made up of two islands including ‘Upolu, where the majority of the population resides, and Savaii (Figure 1). ‘Upolu contains a single urban center, Apia, in which approximately 35,000 residents live, while the rest of the population lives in semi-rural or rural villages. The participants in this study resided in different geographic regions across the island of ‘Upolu. The majority of participants resided in the Apia Urban Area (AUA, urban) and Northwest ‘Upolu (NWU, periurban), while only a handful of participants resided in the Rest of ‘Upolu (ROU, rural) region.

**Figure 1.**
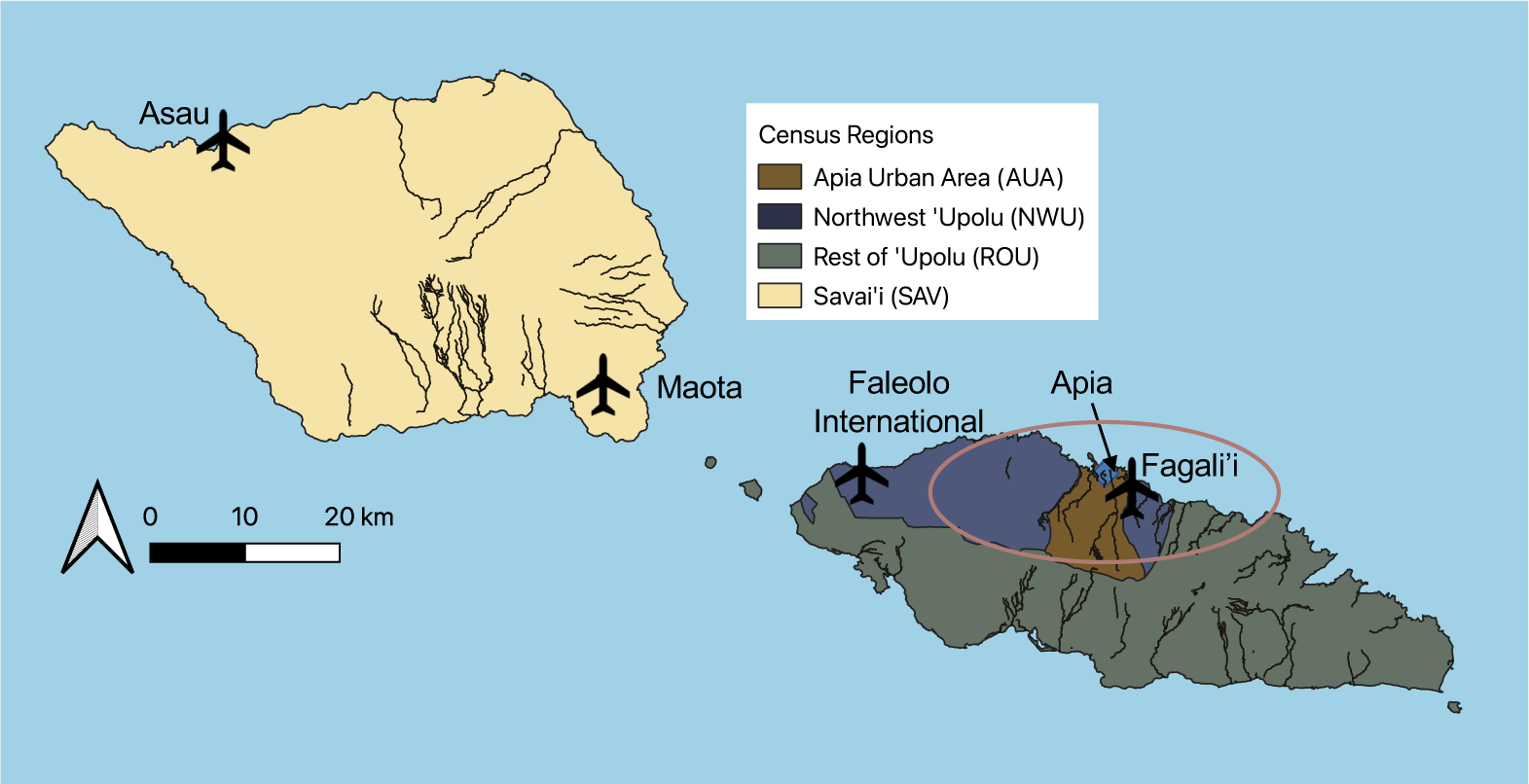
Map depicting census regions and airports of Samoa. Created using QGIS. Data sources: Global Map of Samoa © ISCGM / The Ministry of Natural Resources and Environment, Samoa Ministry of Natural Resources (https://github.com/globalmaps/gmws10, <u>polbnda layer</u>) and NextGIS (Airports, Settlements, and Waterways layers). The circle on the map indicates the approximate area in which research participants were resident.

Parent study inclusion criteria for infants were as follows: their mothers were over 18 years old, 35-41 weeks gestation, had uncomplicated singleton pregnancies, planned to give birth at TTM Hospital, and lived within a 30-minute distance from Apia for easy follow-up. Written informed consent was obtained from all mothers, and the study was approved by Institutional Review Boards at Yale University, the University of Pittsburgh, and the Health Research Committee of the Samoa Ministry of Health. As part of the *Foafoaga o le Ola* study, trained bilingual Samoan research assistants also assessed the mother-infant dyads at 1-week, 2-months, and 4-months post-birth. Data on behavior, environment, and physical measurements were collected, and biospecimens were collected for later analysis. Additional inclusion criteria for this ancillary study included the availability of stored biospecimens from which to quantify PFAS concentrations, focusing on infant cord blood collected at birth and infant DBS collected at 4 months post-birth.

### 2.2 Biospecimen collection

At the pre-birth recruitment visit, the medical charts of women who agreed to provide a cord blood sample were flagged in the medical record. Infant cord blood samples were obtained by trained clinical staff immediately post-birth, with approximately 6 mL collected into a syringe, and 3 mL transferred to a serum separator tube. Standard processing procedures were followed, and serum was stored in cryovials at −80°C. Samples were shipped on dry ice to Yale University for storage, and then to the University of Pittsburgh for PFAS measurement.

At 4 months post-birth, infant capillary blood was collected on a Whatman 903 Protein Saver Card by trained research staff using a heel stick procedure. After cleaning the skin, the blood collection card was placed in contact with the heel to allow the blood to saturate the designated circles. The blood spots were air dried and refrigerated at 4°C, and then shipped at room temperature to Yale University for storage, and then to the University of Pittsburgh for PFAS data collection.

### 2.3 PFAS measurement

We analyzed concentrations (ng/mL) of 40 PFAS analytes (Table S1) in all available cord blood samples (n=66). Quantification was performed based on established methods with some modifications.^21^ Specifically, 100 µL of serum was transferred into a 2-mL polypropylene tube and then spiked with 24 isotopic PFAS (Wellington Laboratories; the list of compounds and the amount spiked can be found in Table S2) as surrogate standards. Next, 250 µL of 0.1 M formic acid and 750 µL of methanol were added to each tube, followed by vortexing for 10 s and then sonication for 10 min. The supernatant was transferred into a new tube and 750 µL of methanol was added into the sample tube for a second round of extraction. The combined supernatants were concentrated to 100 µL, transferred into LC vials, spiked with injection internal standards of 7 isotopic PFAS (Wellington Laboratories; Table S2), and stored at 4℃ for instrumental analysis. PFAS quantification was performed based on methods described in US EPA Draft 2 Method 1633^22^ using a Thermo Scientific TSQ Quantum triple quadrupole mass spectrometer (LC-MS/MS).

Method validation was performed prior to analysis of participant samples by spiking native PFAS (ranging from 0.2 to 2 ng depending on the analyte, Table S1) and isotopic PFAS (Table S2) as surrogates into deionized water; recoveries ranged from 62.8% to 111%. The limits of detections (LOD) for PFAS were determined as the lowest concentrations that meet a signal-to-noise ratio of 3:1 on the instrument and ranged from 0.1 to 2.5 ng/mL (Table S1). Samples were run in batches of approximately 20 samples. Each batch included two laboratory blanks and 1 sample for duplicate. No targeted PFAS were detected in blank samples. The relative standard deviation of duplicated samples ranged from 3.53-22.6%. The isotopic dilution method was used for quantifying PFAS, producing recovery-corrected results.

In an exploratory arm of the study, 50 purposefully selected DBS cards with amble blood content were chosen for PFAS analysis. Small, uniform punches were taken using a 6 mm QIAGEN UniCore Punch Kit (Fisher Scientific) from the center of the blood circles. The hole punch instrument was cleaned between punches to prevent contamination across samples. Based on availability, two to five holes were punched out from a single sample. The sample preparation and instrumental analysis of PFAS were identical to the cord blood samples, except that we were not able to include duplicate samples for DBS data due to limited blood volume. Blood volume was estimated based on the assumption that a single 6 mm punch contained 6.6 microliters of blood.^23^ Specifically, analyte-specific concentrations were calculated by dividing the detected mass by 0.0066 mL multiplied by the number of punches.

### 2.4 Phenotype data

To explore PFAS concentrations across samples, we used phenotype data collected through the *Foafoaga o le Ola* study.^19^ Before birth, a comprehensive set of maternal characteristics was collected through medical record review, questionnaires, and physical assessments. These characteristics included age, years of education, relationship status, census region of residence, and height/weight. Social data included the material lifestyle score, a measure of socioeconomic resources commonly used in Samoa. The score is calculated as the number of 15 possible household assets owned by participants (house, fridge, freezer, stereo, portable speaker, television, VCR/DVD, couch, carpet/rugs, washing machine, landline telephone, mobile phone, computer/laptop, electricity, and motor vehicle).^24, 25^ Infant date of birth and sex were parent-reported at the 1-week assessment. To further describe and characterize the sample, infant birth weight and length were obtained from hospital medical records, and follow up weight and length were measured using standardized instruments—an infant scale (SECA 354, SECA, Hamburg, Germany) and a length board (SECA 417, SECA, Hamburg, Germany). Breastfeeding status was reported by mothers at each follow-up assessment, distinguishing between exclusively breastfed and formula/mixed fed infants (combined because of high rate of exclusive breastfeeding and small sample size).

Of note, dyads were enrolled pre-birth, which resulted in some parents consenting for infant cord blood collection but being lost to follow-up before follow-up visits. Consequently, phenotype data, including infant sex, were missing for some infants. However, to comprehensively characterize PFAS concentrations, data were still collected from these samples.

### 2.5 Statistical analyses

All statistical and graphical descriptions were conducted using R version 4.2.1.^26^ Standard descriptive statistics were computed based on each variable’s level of measurement to provide a comprehensive overview of the data. The 25^th^, 50^th^ (median), 75^th^, and 95^th^ percentiles were reported across PFAS data. Data distributions and patterns were examined both graphically and statistically. Rain cloud or sina with violin plots, created using the ggplot2 R package^27^, were used to describe the data for all participants and subgroups categorized by breastfeeding status (4 months only). Across the majority of analytes, LODs resulted in censored data or “non-detects” below these limits. To visualize data distributions more clearly, values below the LOD were retained on the plots, graphically depicted as LOD/2. Plots were created for all analytes with observations (i.e., >LOD values) for more than a single sample.

We then examined formal differences in cord blood and DBS PFAS concentrations by sex, census region of residence, socioeconomic resources as well as differences in DBS PFAS by nutrition source. In analyzing the censored data, we avoided introducing bias through arbitrary estimates (i.e., replacing non-detect values with the LOD or LOD/2) and instead employed robust approaches that accounted for the underlying distribution of censored data as recommended elsewhere.^28^ Specifically, we used regression using maximum likelihood estimation implemented via the cencorreg function from the NADA2 R package.^29^ Given the data skewness, PFAS data were log transformed. We reported 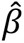, regression estimates for natural log transformed PFAS concentrations, and 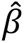 95% confidence intervals. To aid in interpretation, we also reported *exp*(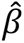), the exponentiated regression estimate, and percent change, calculated as 100 * (*exp*(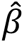) −1). Except for the sex-focused regression, all models controlled for sex. In addition, DBS analyses controlled for infant age in days. In *post hoc* analyses, we added socioeconomic resources as a covariate to the census region and nutrition source models. Because maximum likelihood estimation depends heavily on detections to calibrate the model, these analyses focused on PFAS analytes that were detected in >10% of samples.

Finally, to understand the relationship between PFAS concentrations across time, correlation plots were created using Kendall’s Tau-B and p-values calculated using the ATS function from the NADA2 R package.^29^ These analyses were performed only for analytes detected in >10% of samples across both tissues and accounted for censored data.

Given the exploratory nature of this study, associations with a p-value <0.05 were considered “suggestive”. However, to control the type I error rate, we adjusted for multiple testing by determining the number of effective tests (N_eff_) based on the correlation structure of the PFAS data using the meff() function from the poolr R package.^30^ We then used the Bonferroni correction to compute a threshold for associations considered “significant” by dividing by the number of effective tests (p < 0.05/N_eff_). Associations with a p-value <0.006 or <0.025 were considered “significant” for cord blood and DBS data, respectively.

## 3. RESULTS

### 3.1 Participant characteristics

A flow chart depicting sample availability and selection is presented in Figure S1. Sample characteristics are presented in Table 1 and compared with the parent study in Table S3. The cord blood and DBS samples consisted of 66 (45% female) and 50 (54% female) infant participants, respectively. Only 26 participants overlapped across the two samples. All infants were born at full term (>37 gestational weeks) and mean birth weight ranged from 3.47 kg to 3.60 kg across the two samples. The mean maternal age was approximately 27 years, and over 50% of the sample were residents of the AUA geographic region (Table 1). Characteristics of participants with biospecimens were similar to those without biospecimens (Table S3).

**Table 1.** Participant characteristics.

| Participant Information from Baseline or Birth |  |  |
| --- | --- | --- |
| Characteristic | Cord blood<br>n = 66 | DBS (4 months)<br>n = 50 |
| Infant sex |  |  |
| *Female* | 29 (45%) | 27 (54%) |
| *Male* | 36 (55%) | 23 (46%) |
| *Missing* | 1<sup>a</sup> | 0 |
| Infant birth weight (kg) | 3.60 (0.48) | 3.47 (0.48) |
| *Missing* | 2 | 0 |
| Infant birth length (cm) | 54.73 (5.15) | 52.25 (7.82) |
| *Missing* | 17 | 14 |
| Gestational age at birth (weeks) | 40.08 (0.67) | 39.96 (0.36) |
| *Missing* | 5 | 3 |
| Maternal age, years | 27.33 (5.91) | 27.50 (6.37) |
| *Missing* | 2 | 0 |
| Maternal BMI, kg/m<sup>2</sup> | 35.58 (5.80) | 33.87 (5.35) |
| *Missing* | 7 | 5 |
| Maternal education, years |  |  |
| *Less than primary school* | 2 (3.1%) | 3 (6.0%) |
| *Primary school completed* | 8 (12%) | 5 (10%) |
| *Secondary school/college completed* | 36 (55%) | 29 (58%) |
| *University completed* | 18 (28%) | 13 (26%) |
| *Post-graduate training* | 1 (1.5%) | 0 (0%) |
| *Missing* | 1 | 0 |
| Socioeconomic resources (no. household assets)<sup>b</sup> | 8.1 (2.8) | 8.2 (2.9) |
| *Missing* | 14 | 10 |
| Census region |  |  |
| *AUA* | 39 (60%) | 26 (52%) |
| *NWU* | 26 (40%) | 22 (44%) |
| *ROU* | 0 (0%) | 2 (4.0%) |
| *Missing* | 1 | 0 |
| Infant Data (4 months post-birth) |  |  |
| Characteristic | Cord blood<br>n = 66 | DBS (4 months)<br>n = 50 |
| Age, days | 126.63 (9.72) | 128.98 (16.36) |
| *Missing* | 7 | 1 |
| Nutrition source |  |  |
| *Exclusively breastfed* | 37 (64%) | 35 (74%) |
| *Formula/mixed fed<sup>c</sup>* | 21 (36%) | 12 (26%) |
| *Missing* | 8 | 3 |
| Weight, kg | 7.53 (0.99) | 7.31 (0.94) |
| *Missing* | 7 | 1 |
| Length, cm | 65.06 (2.70) | 64.54 (2.40) |
| *Missing* | 7 | 1 |
| BMI Z score | 0.47 (0.92) | 0.34 (0.85) |
| *Missing* | 7 | 1 |

<sup>a</sup>Lost to follow-up between pre-delivery recruitment and 1-week infant assessment (during which sex is self-reported by parents); <sup>b</sup>Socioeconomic resources based on number of items owned from the 15-item material lifestyle score questionnaire; <sup>c</sup>Formula or mixed-fed categories were combined due to small sample size; n (%) or mean (SD).

### 3.2 Description of PFAS concentrations

PFAS concentrations are summarized in Table 2 and depicted graphically in Figures 2 (cord blood) and 3 (DBS). Of the 40 PFAS analytes tested, 19 were detected in cord blood, with 11 detected in >10% of samples (PFBA, PFPeA, PFHpA, PFOA, PFNA, PFDA, PFUnA, PFTrDA, PFHxS, PFOS, and 9Cl-PF3ONS) while 12 were detected in DBS, with 3 detected in >10% of samples (PFBA, PFHxS, and PFOS). Across cord blood and DBS samples, PFHxS stood out as an analyte detected at high concentrations (median 6.61 and 3.68 ng/mL, respectively). For the DBS data, only PFBA exceeded this with a median concentration of 6.89 ng/mL. Across PFAS classes, perfluoroalkyl carboxylic acids and perfluoroalkyl sulfonic acids were detected at the highest rates. The correlation (Kendall’s Tau or Kendall’s Tau-B) between analytes ranged from −0.18 to 0.57 (cord blood) and 0.06 to 0.49 (DBS) (Figures S2 and S3).

**Figure 2.**
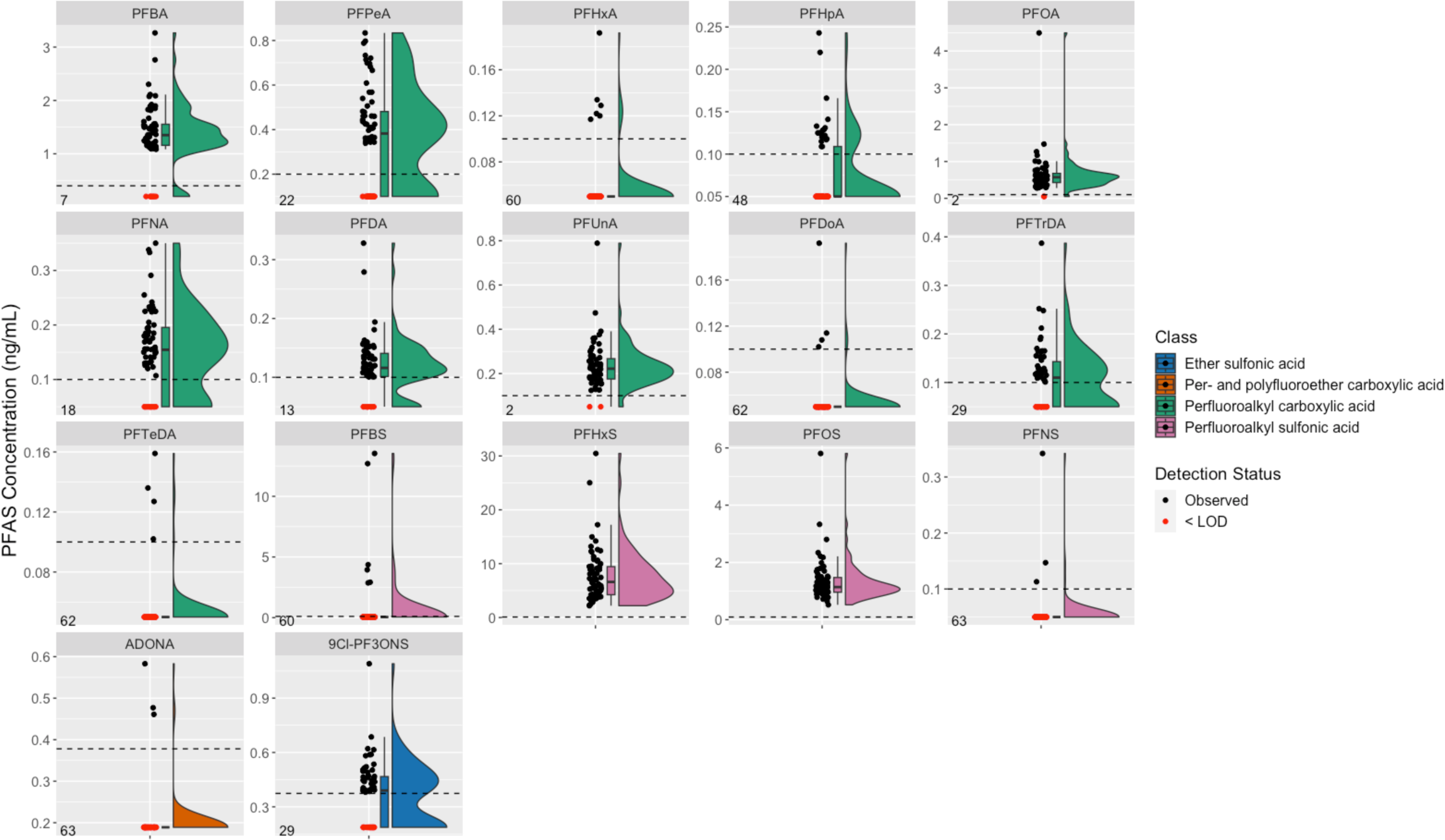
Rain cloud plots describing cord blood PFAS levels for analytes with detectable values (n=66). LOD = limit of detection. Analytes are colored by class; dashed line represents analyte-specific LOD; black dots indicate observed values; red dots indicate < LOD values (depicted graphically as LOD/2); numbers in bottom left corner of plot facets indicate the number of < LOD values. Figure is ordered by class and chain length.

**Table 2.**
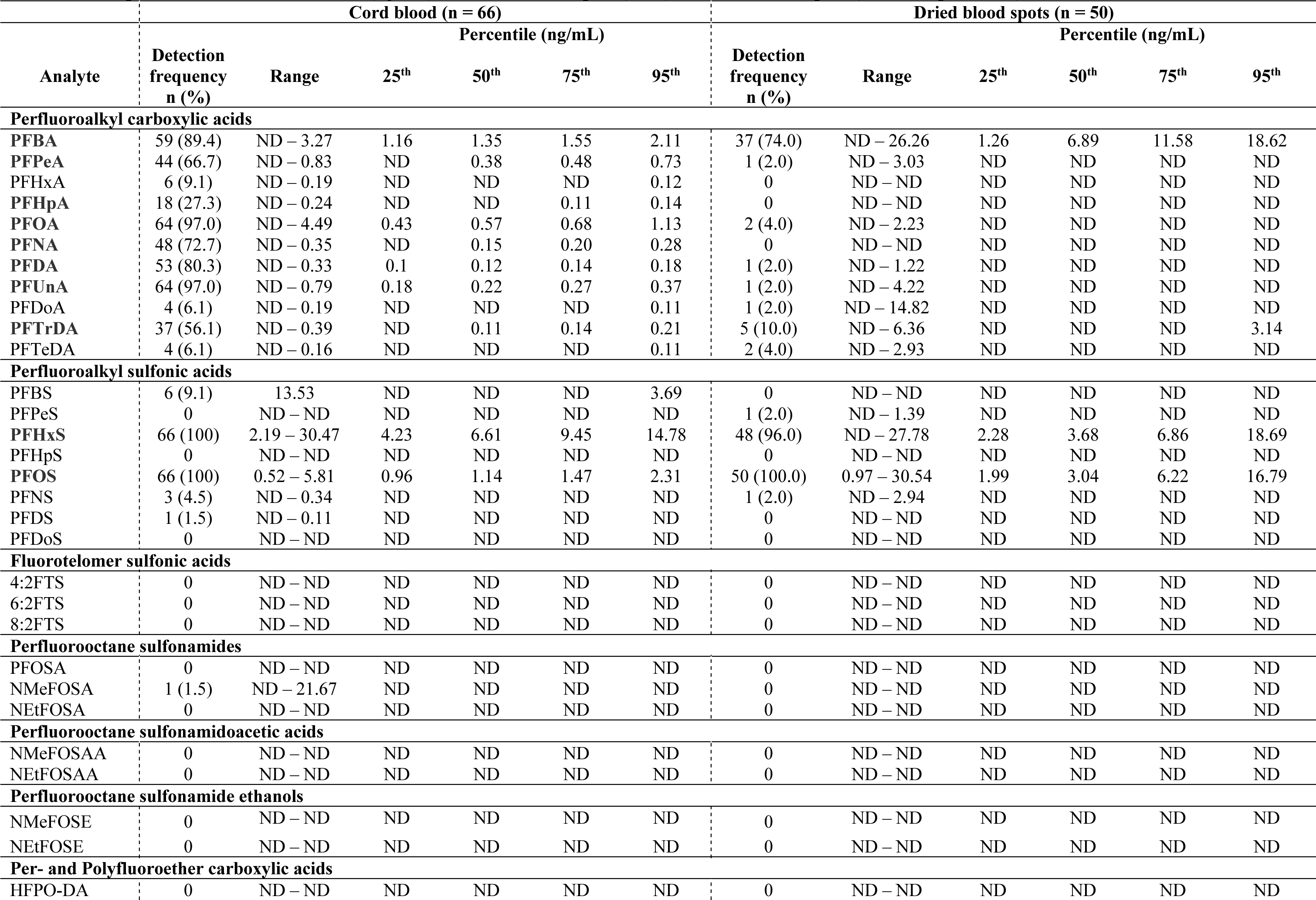
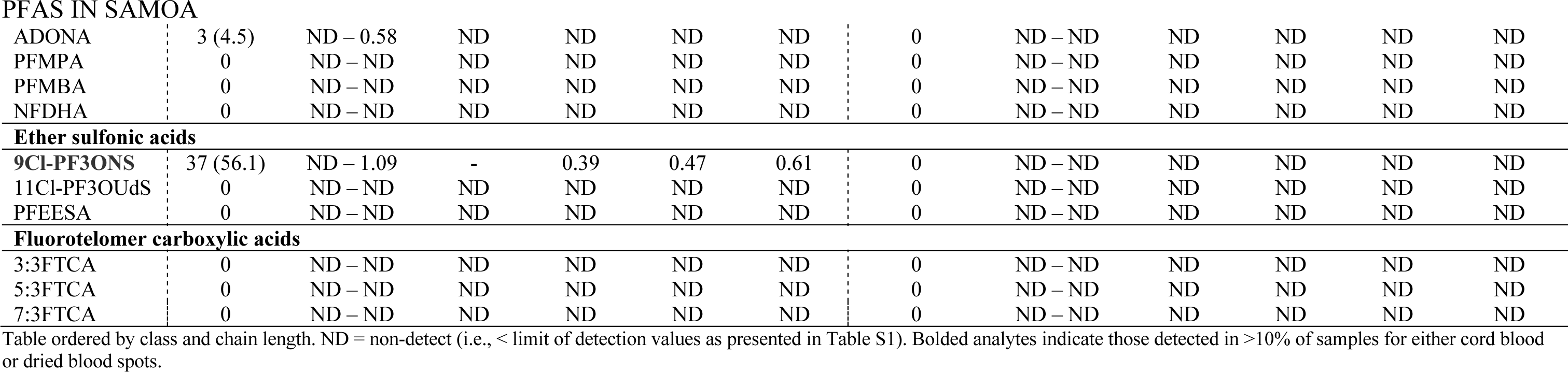
Description of PFAS concentrations (ng/mL) in cord blood samples (birth) and dried blood spots (4 months post-birth).

| Table 2: Description of PFAS concentrations (ng/mL) in cord blood samples (n=66) and dried blood spots (7 months post birth). |  |  |  |  |  |  |  |  |  |  |  |  |
| --- | --- | --- | --- | --- | --- | --- | --- | --- | --- | --- | --- | --- |
| Analyte | Cord blood (n = 66) |  |  |  |  |  | Dried blood spots (n = 50) |  |  |  |  |  |
|  | Detection frequency<br>n (%) | Range | Percentile (ng/mL) |  |  |  | Detection frequency<br>n (%) | Range | Percentile (ng/mL) |  |  |  |
|  |  |  | 25<sup>th</sup> | 50<sup>th</sup> | 75<sup>th</sup> | 95<sup>th</sup> |  |  | 25<sup>th</sup> | 50<sup>th</sup> | 75<sup>th</sup> | 95<sup>th</sup> |
| Perfluoroalkyl carboxylic acids |  |  |  |  |  |  |  |  |  |  |  |  |
| PFBA | 59 (89.4) | ND – 3.27 | 1.16 | 1.35 | 1.55 | 2.11 | 37 (74.0) | ND – 26.26 | 1.26 | 6.89 | 11.58 | 18.62 |
| PFPeA | 44 (66.7) | ND – 0.83 | ND | 0.38 | 0.48 | 0.73 | 1 (2.0) | ND – 3.03 | ND | ND | ND | ND |
| PFHxA | 6 (9.1) | ND – 0.19 | ND | ND | ND | 0.12 | 0 | ND – ND | ND | ND | ND | ND |
| PFHpA | 18 (27.3) | ND – 0.24 | ND | ND | 0.11 | 0.14 | 0 | ND – ND | ND | ND | ND | ND |
| PFOA | 64 (97.0) | ND – 4.49 | 0.43 | 0.57 | 0.68 | 1.13 | 2 (4.0) | ND – 2.23 | ND | ND | ND | ND |
| PFNA | 48 (72.7) | ND – 0.35 | ND | 0.15 | 0.20 | 0.28 | 0 | ND – ND | ND | ND | ND | ND |
| PFDA | 53 (80.3) | ND – 0.33 | 0.1 | 0.12 | 0.14 | 0.18 | 1 (2.0) | ND – 1.22 | ND | ND | ND | ND |
| PFUnA | 64 (97.0) | ND – 0.79 | 0.18 | 0.22 | 0.27 | 0.37 | 1 (2.0) | ND – 4.22 | ND | ND | ND | ND |
| PFDoA | 4 (6.1) | ND – 0.19 | ND | ND | ND | 0.11 | 1 (2.0) | ND – 14.82 | ND | ND | ND | ND |
| PFTTrDA | 37 (56.1) | ND – 0.39 | ND | 0.11 | 0.14 | 0.21 | 5 (10.0) | ND – 6.36 | ND | ND | ND | 3.14 |
| PFTeDA | 4 (6.1) | ND – 0.16 | ND | ND | ND | 0.11 | 2 (4.0) | ND – 2.93 | ND | ND | ND | ND |
| Perfluoroalkyl sulfonic acids |  |  |  |  |  |  |  |  |  |  |  |  |
| PFBS | 6 (9.1) | 13.53 | ND | ND | ND | 3.69 | 0 | ND – ND | ND | ND | ND | ND |
| PFPeS | 0 | ND – ND | ND | ND | ND | ND | 1 (2.0) | ND – 1.39 | ND | ND | ND | ND |
| PFHxS | 66 (100) | 2.19 – 30.47 | 4.23 | 6.61 | 9.45 | 14.78 | 48 (96.0) | ND – 27.78 | 2.28 | 3.68 | 6.86 | 18.69 |
| PFHpS | 0 | ND – ND | ND | ND | ND | ND | 0 | ND – ND | ND | ND | ND | ND |
| PFOS | 66 (100) | 0.52 – 5.81 | 0.96 | 1.14 | 1.47 | 2.31 | 50 (100.0) | 0.97 – 30.54 | 1.99 | 3.04 | 6.22 | 16.79 |
| PFNS | 3 (4.5) | ND – 0.34 | ND | ND | ND | ND | 1 (2.0) | ND – 2.94 | ND | ND | ND | ND |
| PFDS | 1 (1.5) | ND – 0.11 | ND | ND | ND | ND | 0 | ND – ND | ND | ND | ND | ND |
| PFDoS | 0 | ND – ND | ND | ND | ND | ND | 0 | ND – ND | ND | ND | ND | ND |
| Fluorotelomer sulfonic acids |  |  |  |  |  |  |  |  |  |  |  |  |
| 4:2FTS | 0 | ND – ND | ND | ND | ND | ND | 0 | ND – ND | ND | ND | ND | ND |
| 6:2FTS | 0 | ND – ND | ND | ND | ND | ND | 0 | ND – ND | ND | ND | ND | ND |
| 8:2FTS | 0 | ND – ND | ND | ND | ND | ND | 0 | ND – ND | ND | ND | ND | ND |
| Perfluorooctane sulfonamides |  |  |  |  |  |  |  |  |  |  |  |  |
| PFOSA | 0 | ND – ND | ND | ND | ND | ND | 0 | ND – ND | ND | ND | ND | ND |
| NMeFOSA | 1 (1.5) | ND – 21.67 | ND | ND | ND | ND | 0 | ND – ND | ND | ND | ND | ND |
| NEtFOSA | 0 | ND – ND | ND | ND | ND | ND | 0 | ND – ND | ND | ND | ND | ND |
| Perfluorooctane sulfonamidoacetic acids |  |  |  |  |  |  |  |  |  |  |  |  |
| NMeFOSAA | 0 | ND – ND | ND | ND | ND | ND | 0 | ND – ND | ND | ND | ND | ND |
| NEtFOSAA | 0 | ND – ND | ND | ND | ND | ND | 0 | ND – ND | ND | ND | ND | ND |
| Perfluorooctane sulfonamide ethanols |  |  |  |  |  |  |  |  |  |  |  |  |
| NMeFOSE | 0 | ND – ND | ND | ND | ND | ND | 0 | ND – ND | ND | ND | ND | ND |
| NEtFOSE | 0 | ND – ND | ND | ND | ND | ND | 0 | ND – ND | ND | ND | ND | ND |
| Per- and Polyfluoroether carboxylic acids |  |  |  |  |  |  |  |  |  |  |  |  |
| HFPO-DA | 0 | ND – ND | ND | ND | ND | ND | 0 | ND – ND | ND | ND | ND | ND |
| PFAS IN SAMOA |  |  |  |  |  |  |  |  |  |  |  |  |
| ADONA | 3 (4.5) | ND – 0.58 | ND | ND | ND | ND | 0 | ND – ND | ND | ND | ND | ND |
| PFMPA | 0 | ND – ND | ND | ND | ND | ND | 0 | ND – ND | ND | ND | ND | ND |
| PFMBA | 0 | ND – ND | ND | ND | ND | ND | 0 | ND – ND | ND | ND | ND | ND |
| NFDHA | 0 | ND – ND | ND | ND | ND | ND | 0 | ND – ND | ND | ND | ND | ND |
| **Ether sulfonic acids** |  |  |  |  |  |  |  |  |  |  |  |  |
| **9Cl-PF3ONS** | 37 (56.1) | ND – 1.09 | - | 0.39 | 0.47 | 0.61 | 0 | ND – ND | ND | ND | ND | ND |
| 11Cl-PF3OUdS | 0 | ND – ND | ND | ND | ND | ND | 0 | ND – ND | ND | ND | ND | ND |
| PFEESA | 0 | ND – ND | ND | ND | ND | ND | 0 | ND – ND | ND | ND | ND | ND |
| **Fluorotelomer carboxylic acids** |  |  |  |  |  |  |  |  |  |  |  |  |
| 3:3FTCA | 0 | ND – ND | ND | ND | ND | ND | 0 | ND – ND | ND | ND | ND | ND |
| 5:3FTCA | 0 | ND – ND | ND | ND | ND | ND | 0 | ND – ND | ND | ND | ND | ND |
| 7:3FTCA | 0 | ND – ND | ND | ND | ND | ND | 0 | ND – ND | ND | ND | ND | ND |

### 3.3 PFAS concentrations and participant characteristics

To better understand PFAS concentrations by participant characteristics, we examined associations between log transformed PFAS concentrations and sex, census region of residence, and socioeconomic resources (Table 3).

**Table 3.**
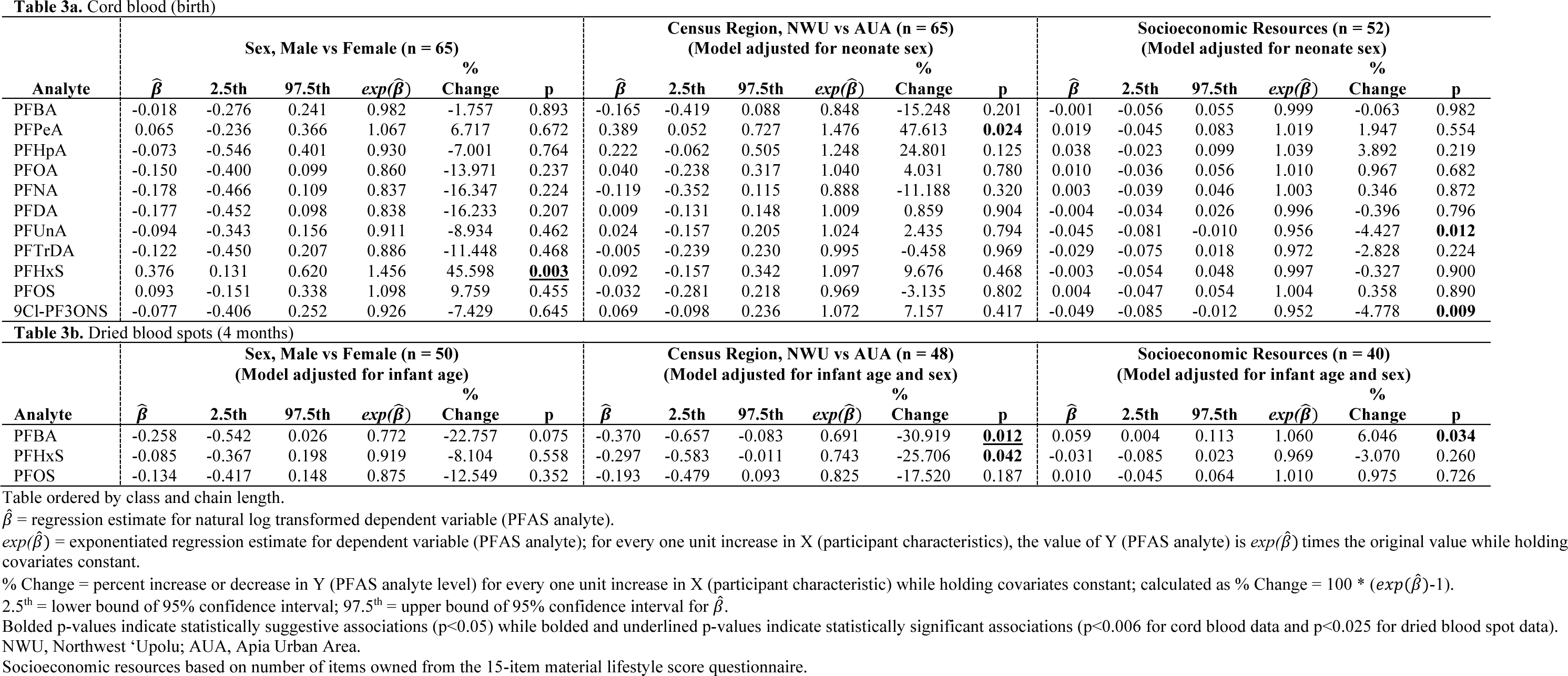
Results of regression using maximum likelihood estimation (MLE) examining associations between natural log-transformed cord blood PFAS levels (dependent variable) and participant factors of sex, census region of residence, and socioeconomic resources.

| Table 3a. Cord blood (birth) |  |  |  |  |  |  |  |  |  |  |  |  |  |  |  |  |  |  |
| --- | --- | --- | --- | --- | --- | --- | --- | --- | --- | --- | --- | --- | --- | --- | --- | --- | --- | --- |
| Analyte | Sex, Male vs Female (n = 65) |  |  |  |  |  | Census Region, NWU vs AUA (n = 65) |  |  |  |  |  | Socioeconomic Resources (n = 52) |  |  |  |  |  |
| | $\hat{\beta}$ | 2.5th | 97.5th | $exp(\hat{\beta})$ | % Change | p | $\hat{\beta}$ | 2.5th | 97.5th | $exp(\hat{\beta})$ | % Change | p | $\hat{\beta}$ | 2.5th | 97.5th | $exp(\hat{\beta})$ | % Change | p |
| PFBA | -0.018 | -0.276 | 0.241 | 0.982 | -1.757 | 0.893 | -0.165 | -0.419 | 0.088 | 0.848 | -15.248 | 0.201 | -0.001 | -0.056 | 0.055 | 0.999 | -0.063 | 0.982 |
| PFPeA | 0.065 | -0.236 | 0.366 | 1.067 | 6.717 | 0.672 | 0.389 | 0.052 | 0.727 | 1.476 | 47.613 | **0.024** | 0.019 | -0.045 | 0.083 | 1.019 | 1.947 | 0.554 |
| PFHpA | -0.073 | -0.546 | 0.401 | 0.930 | -7.001 | 0.764 | 0.222 | -0.062 | 0.505 | 1.248 | 24.801 | 0.125 | 0.038 | -0.023 | 0.099 | 1.039 | 3.892 | 0.219 |
| PFOA | -0.150 | -0.400 | 0.099 | 0.860 | -13.971 | 0.237 | 0.040 | -0.238 | 0.317 | 1.040 | 4.031 | 0.780 | 0.010 | -0.036 | 0.056 | 1.010 | 0.967 | 0.682 |
| PFNA | -0.178 | -0.466 | 0.109 | 0.837 | -16.347 | 0.224 | -0.119 | -0.352 | 0.115 | 0.888 | -11.188 | 0.320 | 0.003 | -0.039 | 0.046 | 1.003 | 0.346 | 0.872 |
| PFDA | -0.177 | -0.452 | 0.098 | 0.838 | -16.233 | 0.207 | 0.009 | -0.131 | 0.148 | 1.009 | 0.859 | 0.904 | -0.004 | -0.034 | 0.026 | 0.996 | -0.396 | 0.796 |
| PFUnA | -0.094 | -0.343 | 0.156 | 0.911 | -8.934 | 0.462 | 0.024 | -0.157 | 0.205 | 1.024 | 2.435 | 0.794 | -0.045 | -0.081 | -0.010 | 0.956 | -4.427 | **0.012** |
| PFTTrDA | -0.122 | -0.450 | 0.207 | 0.886 | -11.448 | 0.468 | -0.005 | -0.239 | 0.230 | 0.995 | -0.458 | 0.969 | -0.029 | -0.075 | 0.018 | 0.972 | -2.828 | 0.224 |
| PFHxS | 0.376 | 0.131 | 0.620 | 1.456 | 45.598 | **0.003** | 0.092 | -0.157 | 0.342 | 1.097 | 9.676 | 0.468 | -0.003 | -0.054 | 0.048 | 0.997 | -0.327 | 0.900 |
| PFOS | 0.093 | -0.151 | 0.338 | 1.098 | 9.759 | 0.455 | -0.032 | -0.281 | 0.218 | 0.969 | -3.135 | 0.802 | 0.004 | -0.047 | 0.054 | 1.004 | 0.358 | 0.890 |
| 9Cl-PF3ONS | -0.077 | -0.406 | 0.252 | 0.926 | -7.429 | 0.645 | 0.069 | -0.098 | 0.236 | 1.072 | 7.157 | 0.417 | -0.049 | -0.085 | -0.012 | 0.952 | -4.778 | **0.009** |
| Table 3b. Dried blood spots (4 months) |  |  |  |  |  |  |  |  |  |  |  |  |  |  |  |  |  |  |
| Analyte | Sex, Male vs Female (n = 50) |  |  |  |  |  | Census Region, NWU vs AUA (n = 48) |  |  |  |  |  | Socioeconomic Resources (n = 40) |  |  |  |  |  |
| | $\hat{\beta}$ | 2.5th | 97.5th | $exp(\hat{\beta})$ | % Change | p | $\hat{\beta}$ | 2.5th | 97.5th | $exp(\hat{\beta})$ | % Change | p | $\hat{\beta}$ | 2.5th | 97.5th | $exp(\hat{\beta})$ | % Change | p |
| PFBA | -0.258 | -0.542 | 0.026 | 0.772 | -22.757 | 0.075 | -0.370 | -0.657 | -0.083 | 0.691 | -30.919 | **0.012** | 0.059 | 0.004 | 0.113 | 1.060 | 6.046 | **0.034** |
| PFHxS | -0.085 | -0.367 | 0.198 | 0.919 | -8.104 | 0.558 | -0.297 | -0.583 | -0.011 | 0.743 | -25.706 | **0.042** | -0.031 | -0.085 | 0.023 | 0.969 | -3.070 | 0.260 |
| PFOS | -0.134 | -0.417 | 0.148 | 0.875 | -12.549 | 0.352 | -0.193 | -0.479 | 0.093 | 0.825 | -17.520 | 0.187 | 0.010 | -0.045 | 0.064 | 1.010 | 0.975 | 0.726 |

Table ordered by class and chain length.
$\hat{\beta}$ = regression estimate for natural log transformed dependent variable (PFAS analyte).
$exp(\hat{\beta})$ = exponentiated regression estimate for dependent variable (PFAS analyte); for every one unit increase in X (participant characteristics), the value of Y (PFAS analyte) is $exp(\hat{\beta})$ times the original value while holding covariates constant.
% Change = percent increase or decrease in Y (PFAS analyte level) for every one unit increase in X (participant characteristic) while holding covariates constant; calculated as % Change = $100 * (exp(\hat{\beta}) - 1)$ .
2.5<sup>th</sup> = lower bound of 95% confidence interval; 97.5<sup>th</sup> = upper bound of 95% confidence interval for $\hat{\beta}$ .
Bolded p-values indicate statistically suggestive associations ( $p < 0.05$ ) while bolded and underlined p-values indicate statistically significant associations ( $p < 0.006$ for cord blood data and $p < 0.025$ for dried blood spot data).
NWU, Northwest 'Upolu; AUA, Apia Urban Area.
Socioeconomic resources based on number of items owned from the 15-item material lifestyle score questionnaire.

For the cord blood data (Table 3a), we observed an association between natural log transformed PFHxS and sex, with higher concentrations in male infants compared with female infants (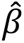 = 0.376 [0.131 to 0.620], p = 0.003). While controlling for neonate sex, we also observed an association between natural log transformed PFPeA and census region of residence, with higher concentrations in infants residing in NWU compared to AUA (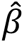 = 0.389 [0.052 to 0.727], p = 0.024). Finally, we observed associations between greater socioeconomic resources and lower log transformed PFUnA (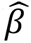 = −0.045 [−0.081 to −0.010], p = 0.012) and 9Cl-PF3ONS (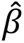 = −0.049 [−0.085 to −0.012], p=0.009) while controlling for sex.

For the DBS data (Table 3b), no statistically significant differences were observed between PFAS concentrations and sex while controlling for infant age. However, we observed associations between census region of residence and natural log transformed PFBA (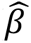 = −0.370 [−0.657 to −0.083], p = 0.012) and PFHxS (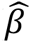 = −0.297 [−0.583 to −0.011], p = 0.042) while controlling for infant age and sex, with lower PFAS concentrations observed in NWU compared with AUA. However, these geographic associations did not persist when controlling for socioeconomic resources (Table S4). We also observed an association between higher natural log transformed PFBA and greater socioeconomic resources while controlling for infant age and sex (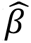 = 0.059 [0.004 to 0.113], p = 0.034).

Lastly, we examined associations between DBS PFAS concentration and nutrition source at four months of life (Table 4 and Figure S4). We observed associations between nutrition source and natural log transformed PFBA (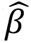 = 0.601 [0.253 to 0.949], p = 0.001) and PFHxS (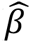 = 0.468 [0.121 to 0.815], p = 0.008) while controlling for infant age and sex, with higher PFAS concentrations observed in formula- or mixed-fed infants compared to exclusively breastfed infants. These associations persisted even after controlling for socioeconomic resources (Table S5).

**Table 4.**
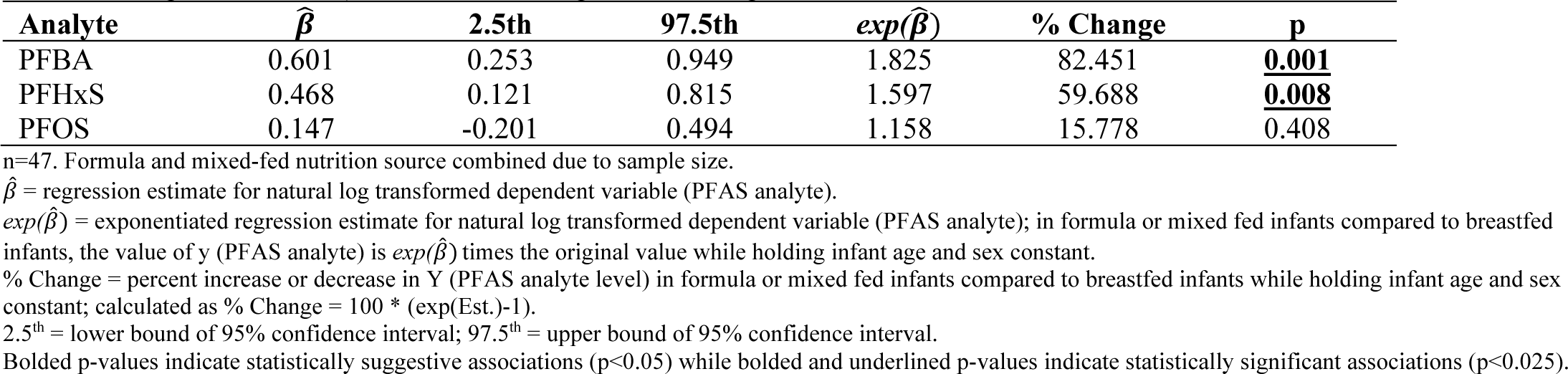
Results of regression using maximum likelihood estimation (MLE) examining associations between natural log-transformed cord blood PFAS levels (dependent variable) and nutrition source (formula-fed or mixed-fed vs. exclusive breastfeeding as reference) while controlling for infant age and sex.

Finally, as shown in Figure 4, we examined correlation between PFAS analytes that were detected in >10% of samples across both tissues (PFBA, PFHxS, and PFOS, n = 26). A statistically significant correlation was observed only for PFHxS (Kendall’s Tau-B = 0.29, p = 0.037).

**Figure 3.**
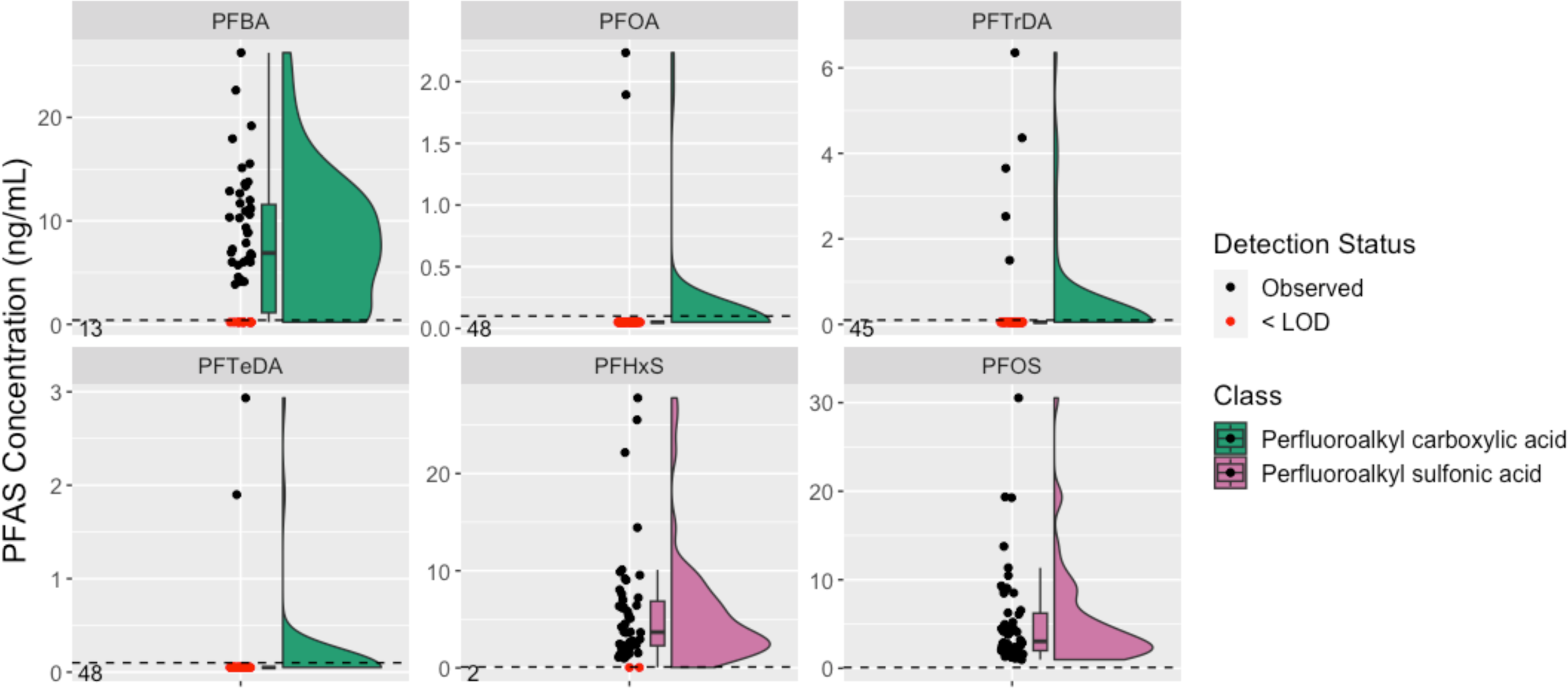
Rain cloud plots describing dried blood spot (4 months) PFAS levels for analytes with detectable values (n=50). LOD = limit of detection. Analytes are colored by class; dashed line represents analyte-specific LOD; black dots indicate observed values; red dots indicate < LOD values (depicted graphically as LOD/2); numbers in bottom left corner of plot facets indicate the number of < LOD values. Figure is ordered by class and chain length.

**Figure 4.**
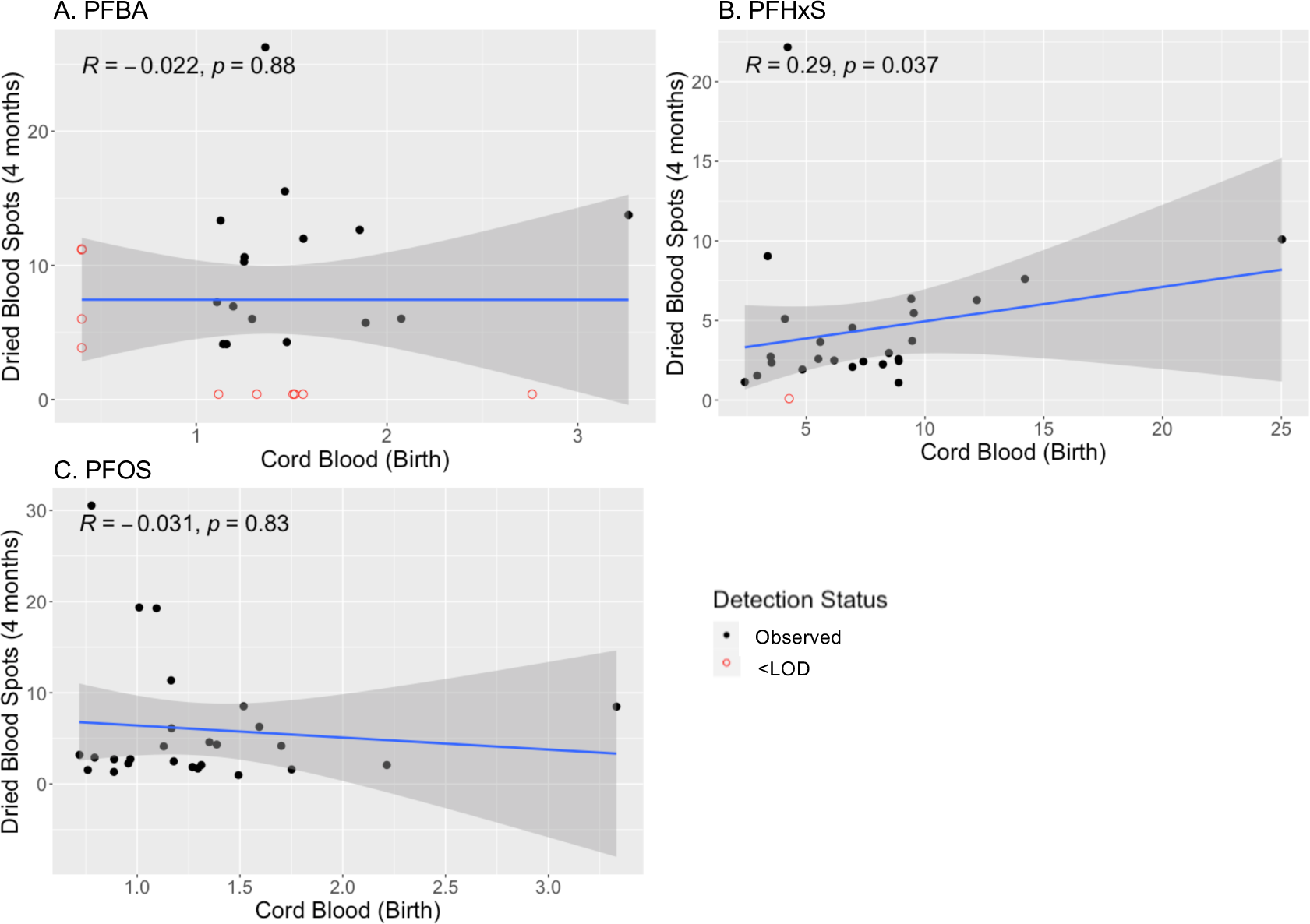
Correlation between PFAS levels in cord blood (birth) and dried blood spots (4 months). N = 26. LOD = limit of detection. Plots depict associations between raw PFAS levels (i.e., untransformed) detected in cord blood (birth) and dried blood spots (4 months) for analytes detected in both tissues (A. PFBA, B. PFHxS, and C. PFOS). Black, solid circles depict observed values while red, open circles depict non-detect values (i.e., those <LOD). R represents Kendall’s Tau (plot C) or Kendall’s Tau-B (plots A and B).

## 4. DISCUSSION

### 4.1 Comparison with existing literature

We present the first data on PFAS contamination in Samoa. Among the detected analytes, PFOS, PFHxS, and PFBA stood out as commonly detected analytes in both cord blood and DBS, which aligns with findings in other studies on PFAS.^6^ Notably, PFOS was present in all samples while PFHxS was observed in 100% of cord blood samples and 96% of DBS. Given the wide application of these chemicals both nationally and globally^2^, and detection of these analytes in blood of 97% of USA residents surveyed in 2011-2012^31^, these observations are not surprising.

In comparing our results with existing literature, the median concentrations of PFOS detected in this study (1.14 ng/mL in cord blood and 3.04 ng/mL in DBS) were lower than those reported in cord blood in New York, US (6.32 ng/mL; sampled in 2001-2002)^32^ and Michigan, US (5.3 ng/mL; sampled in 2010-2019)^33^ and comparable or higher than those reported in Shanghai, China (2.5 ng/mL; sampled in 2012-2012)^34^ and Belgium (0.99 ng/mL; sampled in 2008-2014)^35^.

Interestingly, relatively higher concentrations of PFHxS were detected in this study (median 6.61 ng/mL in cord blood and 3.68 ng/mL in DBS) compared to those reported in cord blood in existing literature (i.e., 3.2 ng/mL in Michigan, US^33^; 0.66 ng/mL in New York, US^32^; 0.15 ng/mL in Belgium^35^; 0.18 ng/mL in Shanghai, China^34^). The high PFHxS concentrations could reflect a replacement transition after the phase-out of PFOS and PFOA since 2000s.^15^ While PFHxS has also been regulated in the US, and some studies have reported serum PFHxS concentrations to be gradually decreasing over the past two decades^36^, this long chain PFAS continues to be detected at high rates in the Pacific, including Vanuatu^15^ and now Samoa. This observation could be a result of continued import of products containing PFHxS from other counties that have no restriction on PFHxS. Additionally, long biological half-lives of PFHxS could lead to its bioaccumulation and presence in the blood for years.

PFBA, used as an additional replacement of PFOA in the USA^37^, was detected with comparable or slightly higher levels than PFOA in this study, indicating the widespread use of and exposure to PFAS alternatives. PFBA is a short-chain PFAS that has not received a great deal of regulatory scrutiny as it is rapidly excreted, despite its association with thyroid hormone levels in humans^38^ and systemic toxicity in a murine model.^37^

Other analytes, including PFPeA, PFOA, PFNA, PFDA, PFUnA, PFTrDA, and 9Cl-PF3ONS, were also detected in over 50% of cord blood samples but at generally lower concentrations compared to existing literature where comparisons existed.^32, 34, 39^ Many of these are long-chain PFAS with a tendency to bioaccumulate. These findings, paired with evidence of PFAS contamination across the Asia Pacific region^15, 40, 41^ and recent governmental efforts to modernize water infrastructure and reduce PFAS contamination in neighboring American Samoa^42^ highlight the importance of monitoring PFAS across Samoa.

### 4.2 Associations with participant factors

In further characterizing the PFAS concentrations across participant characteristics, we observed an association between higher PFHxS and male sex in cord blood. This finding is inconsistent with a 2012-2013 study of cord blood PFAS levels from Shanghai which identified no differences in PFAS concentrations by sex.^34^ Other literature, however, suggests that there are sex differences in specific analytes, including PFHxS, at older ages^43–45^ and outcome-impacting sex-specific associations between PFAS and DNA methylation^33^, an epigenetic regulator of gene expression. No significant sex differences were observed in PFAS concentrations of DBS collected 4 months post-birth, however. Additional work is needed to better understand potential differences in PFAS bioaccumulation by sex.

In terms of geographic region, we hypothesized that we would observe higher concentrations of PFAS in infants residing in NWU (which is home to ‘Upolu’s two airports) compared with the AUA. In cord blood, only a single analyte, PFPeA, was associated with geographic region in our hypothesized direction. In the DBS data, two analytes, PFHxS and PFBA were associated with geographic region, but in the opposite direction, with lower levels observed in NWU compared to AUA. In trying to better understand these associations across geographic region, *post hoc* analyses controlling for socioeconomic resources washed out associations of geographic region and PFHxS and PFBA in DBS, while the association with PFPeA in cord blood remained.

In high-income countries, while chronic exposure to toxicants has been historically observed at higher rates in lower income communities that are often located closer to exposure sources^46^, the literature suggests that concentrations of PFAS generally increase with socioeconomic status indicators such as income, which is mainly attributed to discordant use of consumer products or dietary differences.^47, 48^ Samoa is a lower-middle income country, and our prior work has revealed that there are often unexpected directions of effect, particularly related to social variables.^49^ In fact, we observed lower levels of cord blood PFUnA in neonates whose mothers had greater socioeconomic resources. It is possible that this observation is a result of the ability of families with greater socioeconomic resources to purchase fresh whole foods, compared with lower income families who must rely on cheaper processed and packaged foods. Similar to geographic region, however, effect directions differed across tissues with higher levels of DBS PFBA in individuals with greater socioeconomic resources 4 months post-birth. Additional work in larger sample sizes is needed to better understand these associations.

Finally, we observed higher DBS concentrations of two PFAS analytes, PFHxS and PFBA, in infants who were formula- or mixed-fed at 4 months post-birth compared to infants who were exclusively breastfed. These strong associations persisted even after controlling for socioeconomic resources. These findings are inconsistent with prior work from the Faroe Islands that found, with the exception of PFHxS, PFAS concentrations generally increased at higher rates in breastfed infants compared to formula-fed infants, though they did not investigate exposure sources at length.^50^ While the levels of PFAS in formula are generally low^51^, there has been a call for more research on this topic.^52^ In Samoa, the majority of formula is powder and reconstitution with water is required. Given the need for improved water infrastructure, it is possible that the higher concentrations of PFAS observed in formula/mixed-fed infants could be related to water contamination. Additional work is needed to understand PFAS risk from human milk or formula in this setting, particularly across formula types, as modifiable nutrition-related interventions offers promise to reduce PFAS exposure or bioaccumulation.^53^

### 4.3 Potential sources of PFAS contamination in Samoa

Sources of PFAS exposure are generally consistent with some variations based on the specific compound, use, environmental behavior, and potential for bioavailability and bioaccumulation. Without conducting environmental monitoring studies, however, we cannot say with confidence what the main exposure sources of PFAS are in Samoa. Possible routes of PFAS exposure include contaminated food (e.g., ocean fish, canned foods, meat, dairy) or drinking water^54, 55^, with added potential transfer to infants through breastfeeding or formula preparation. While water treatment plants and national water standards exist across Samoa, not all families rely on public water sources and those who are dependent on rainfall and surface water may have additional potential exposure routes.^56^ Other routes of potential exposure include soil contact, inhalation (e.g., air, dust), or consumer products via dermal exposure.^54, 55^ For example, PFAS have been detected in consumer products targeting infants and children (e.g., clothing, bedding, bibs), often lacking explicit disclosure of chemical additives.^57, 58^ Additional work is needed to survey Samoan individuals regarding types of consumer products that they use frequently, and perform environmental and product sampling.

### 4.3. Limitations

Despite the study’s strengths, including its longitudinal design and examination of 40 PFAS analytes across two tissues in a unique sample of infants from Samoa, there are limitations that should be acknowledged. First, given the exploratory nature of this study and challenges of collecting biospecimens in a low-resource setting like Samoa, the sample size was small; future investigations in larger sample sizes are needed with supplemental investigations of environmental samples. Further, while DBS represent an important public health monitoring tool, particularly for low-resource settings, a major challenge of this tissue centers on blood volume uncertainty across punches as a product of discordant card saturation, blood viscosity, or pressure applied during punching. The PFAS concentrations in DBS were generally lower than that detected in cord blood, which were possibly a result of tissue type/blood volume. While we strategically adjusted for blood volume^23^, future directions include comparing PFAS levels across peripheral blood/DBS collected from the same individual, as well as comparing our blood volume normalization approach with more robust methods such as measuring specific gravity or hemoglobin content of samples.

## 5. Conclusion

In conclusion, this study presents the first evidence of PFAS contamination in Samoa and underscores a broader global issue of environmental justice across communities that face higher relative risks but have limited resources and regulatory frameworks. Expanded investigations of PFAS contamination across environmental compartments and residents of Samoa are needed, including examination of associations of PFAS concentrations and health outcomes.

Understanding determinants of PFAS concentrations and their effects is crucial for exposure estimation and implementing effective public health interventions.

## DECLARATIONS

## Acknowledgements

We would like to thank the participants for their involvement in this research as well as the Samoa Ministry of Health, the Samoa National Health Service, and the antenatal and delivery room nurses and clinicians at TTM Hospital for their support of this work. A particular fa’afetai tele lava to Theresa Atanoa, Madison Rodman, and Elise Claffey and OLaGA Program Coordinator Alysa Pomer. We would also like to extend our appreciation to: Dr. Dennis Helsel for his work in analytical approaches for handling censored environmental data and the many training courses that he has made available; the instructors and organizers of the Columbia University SHARP GIS Workshop for creating a training course related to mapping; and Dr. Michael Sikorski for his counsel in creating Figure 1 which depicts the Samoa census regions.

## Funding

Research reported in this publication was supported by the National Institutes of Health under award numbers TL1TR001858, and K99HD107030, and L40ES033405 (LWH); the Heilbrunn Family Center for Nursing Research at the Rockefeller University (LWH); and the National Science Foundation under award number 1749911 (KJA). Infrastructural support was provided by R0HL1093093 (Stephen T McGarvey). The content is solely the responsibility of the authors and does not necessarily represent the official views of the supporting foundations.

## Conflict of Interest

The funders had no role in the design of the study; collection, analyses, or interpretation of data; writing of the manuscript; or decision to publish the results. As such, the authors declare no conflict of interest.

## Ethical Approval

This study was approved by the Institutional Review Boards (IRB) at Yale University (2000021076). Data analysis activities at the University of Pittsburgh were determined to be exempt (STUDY20020099) based on the receipt of only deidentified data. The study was also approved by the Health Research Committee of the Samoa Ministry of Health.

## Consent to Participate

Written informed consent was obtained from all participant guardians prior to enrollment.

## Availability of Data

The participants included in this study were not consented for data sharing. However, we recognize the importance of data sharing for scientific advancement and to ensure transparency and reproducibility of results and are therefore committed to providing detailed and aggregated results of analyses, or organizing partnerships that allow data sharing with approval from the Samoa Health Research Committee. Researchers interested in discussing this in more detail can contact the corresponding author at for more information.

## Authors’ Roles

Heinsberg, Weeks, Conley, Ng, and Hawley conceived and designed the present study. Arslanian and Hawley were the PIs of the parent study and responsible for procurement of parent study funding, cohort recruitment, and leading phenotype data and biospecimen collection. Heinsberg was the PI of this PFAS-focused study and was responsible for acquisition of the study funding. Niu, Chen, and Bedhi, with supervision from Ng, were responsible for generating the PFAS data. Heinsberg was responsible for biospecimen handling with oversight from Conley. Heinsberg performed all statistical analyses with oversight from Weeks. Heinsberg and Niu were responsible for the first draft of the manuscript and critically revised based on co-author feedback. All authors contributed to the interpretation of the data and results. All authors reviewed, critically revised, and approved the final manuscript. All authors agree to be accountable for all aspects of the work.

## Notes

### Competing Interest Statement

The authors have declared no competing interest.

