## Supplementary Information for "Characterization of Per- and Polyfluoroalkyl Substance (PFAS) concentrations in a community-based sample of infants from Samoa"

*Supplementary Material, Additional File 1*

Lacey W. Heinsberg, PhD, RN*^; Shan Niu, PhD*; Kendall J. Arslanian, PhD; Ruiwen Chen, PhD; Megha Bedi, PhD; Folla Unasa-Apelu; Ulai T. Fidow, MBBS, MMed; Christina Soti-Ulberg, MA; Yvette P. Conley, PhD, FAAN; Daniel E. Weeks, PhD; Carla A. Ng, PhD; Nicola L. Hawley, PhD

*Equal contributions as first author
^Corresponding author
Department of Human Genetics
Graduate School of Public Health
University of Pittsburgh


**TABLE OF CONTENTS**


| Table/Figure | Title | Page |
|---|---|---|
| Abstract | Translated Abstract (Samoan) | 2 |
| Table S1 | PFAS analytes, classes, limits of detection (LOD), and amounts of spiked PFAS for the method validation. | 3 |
| Table S2 | Extracted internal standards and injection internal standards used for PFAS quantification. | 5 |
| Figure S1 | Sample flow chart. | 6 |
| Table S3 | Expanded sample characteristics with full cohort and subsample comparisons. | 7 |
| Figure S2 | Heat map depicting correlation structure of cord blood PFAS data (n = 66). | 8 |
| Figure S3 | Heatmap depicting correlation structure of dried blood spot (4 months) PFAS data (n = 50). | 9 |
| Table S4 | Results of regression using maximum likelihood estimation (MLE) examining associations between natural log-transformed cord blood PFAS levels (dependent variable) and census region (NWU vs. AUA) while controlling for infant age, sex, *and socioeconomic status*. | 10 |
| Figure S4 | Sina with violin plots describing dried blood spot (4 months) PFAS levels by breastfeeding status at 4 months post-birth (n = 47). | 12 |
| Table S5 | Results of regression using maximum likelihood estimation (MLE) examining associations between natural log-transformed cord blood PFAS levels (dependent variable) and breastfeeding status (formula-fed or mixed-fed vs. exclusive breastfeeding as reference) while controlling for infant age and sex (Table S4a) and infant age, sex, and socioeconomic resources (Table S4b). | 13 |
| Abbreviations | Abbreviations | 14 |

### Translated Abstract

*A practice of our research group is to make an abstract that has been translated to Samoan available. At this time, only English language text is allowed by medRxiv. A Samoan translation is available upon request from.*

**Table S1.** PFAS analytes, classes, limits of detection (LOD), and amounts of spiked PFAS for the method validation.

| Class[1] | Analyte | Name | CAS number | Chain length[2] | LOD[3] (ng/mL) | Amount spike (ng) |
|---|---|---|---|---|---|---|
| Perfluoroalkyl carboxylic acids (PFCAs) | PFBA | Perfluorobutanoic acid | 375-22-4 | C4, Short | 0.40 | 0.8 |
| | PFPeA | Perfluoropentanoic acid | 2706-90-3 | C5, Short | 0.20 | 0.4 |
| | PFHxA | Perfluorohexanoic acid | 307-24-4 | C6, Short | 0.10 | 0.2 |
| | PFHpA | Perfluoroheptanoic acid | 375-85-9 | C7, Short | 0.10 | 0.2 |
| | PFOA | Perfluorooctanoic acid | 335-67-1 | C8, Long | 0.10 | 0.2 |
| | PFNA | Perfluorononanoic acid | 375-95-1 | C9, Long | 0.10 | 0.2 |
| | PFDA | Perfluorodecanoic acid | 335-76-2 | C10, Long | 0.10 | 0.2 |
| | PFUnA | Perfluoroundecanoic acid | 2058-94-8 | C11, Long | 0.10 | 0.2 |
| | PFDoA | Perfluorododecanoic acid | 307-55-1 | C12, Long | 0.10 | 0.2 |
| | PFTrDA | Perfluorotridecanoic acid | 72629-94-8 | C13, Long | 0.10 | 0.2 |
| | PFTeDA | Perfluorotetradecanoic acid | 376-06-7 | C14, Long | 0.10 | 0.2 |
| Perfluoroalkyl sulfonic acids (PFSAs) | PFBS | Perfluorobutanesulfonic acid | 375-73-5 | C4, Short | 0.10 | 0.2 |
| | PFPeS | Perfluoropentanesulfonic acid | 2706-91-4 | C5, Short | 0.10 | 0.2 |
| | PFHxS | Perfluorohexanesulfonic acid | 355-46-4 | C6, Long | 0.10 | 0.2 |
| | PFHpS | Perfluoroheptanesulfonic acid | 375-92-8 | C7, Long | 0.10 | 0.2 |
| | PFOS | Perfluorooctanesulfonic acid | 1763-23-1 | C8, Long | 0.10 | 0.2 |
| | PFNS | Perfluorononanesulfonic acid | 68259-12-1 | C9, Long | 0.10 | 0.2 |
| | PFDS | Perfluorodecanesulfonic acid | 335-77-3 | C10, Long | 0.10 | 0.2 |
| | PFDoS | Perfluorododecanesulfonic acid | 79780-39-5 | C12, Long | 0.10 | 0.2 |
| Fluorotelomer sulfonic acids | 4:2FTS | 4:2 Fluorotelomer sulfonic acid | 757124-72-4 | C6, Short | 0.40 | 0.8 |
| | 6:2FTS | 6:2 Fluorotelomer sulfonic acid | 27619-97-2 | C8, Short | 0.40 | 0.8 |
| | 8:2FTS | 8:2 Fluorotelomer sulfonic acid | 39108-34-4 | C10, Long | 0.40 | 0.8 |
| Perfluorooctane sulfonamides | PFOSA | Perfluorooctanesulfonamide | 754-91-6 | C8, Long | 0.10 | 0.2 |
| | NMeFOSA | N-Methylperfluorooctanesulfonamide | 31506-32-8 | C9, Long | 0.10 | 0.2 |
| | NEtFOSA | N-Ethylperfluorooctanesulfonamide | 4151-50-2 | C10, Long | 0.10 | 0.2 |
| Perfluorooctane sulfonamidoacetic acids | NMeFOSAA | N-Methylperfluorooctanesulfonamidoacetate | 2355-31-9 | C11, Long | 0.10 | 0.2 |
| | NEtFOSAA | N-Ethylperfluorooctanesulfonamidoacetate | 2991-50-6 | C12, Long | 0.10 | 0.2 |
| Perfluorooctane sulfonamide ethanols | NMeFOSE | N-Methyl perfluorooctanesulfonamidoethanesulfonate | 24448-09-7 | C11, Long | 1.0 | 2 |
| | NEtFOSE | N-Ethyl perfluorooctanesulfonamidoethanesulfonate | 1691-99-2 | C12, Long | 1.0 | 2 |
| Per- and polyfluoroether carboxylic acids | PFMPA | Perfluoro-3-methoxypropanoic acid | 377-73-1 | C4, Short | 0.20 | 0.8 |
| | PFMBA | Perfluoro-4-methoxybutanoic acid | 863090-89-5 | C5, Short | 0.20 | 0.8 |
| | NFDHA | Nonafluoro-3,6-dioxaheptanoic acid | 151772-58-6 | C5, Short | 0.20 | 0.8 |

| | | | | | | |
|---|---|---|---|---|---|---|
| | HFPO-DA | Hexafluoropropylene oxide dimer acid | 13252-13-6 | C6, Short | 0.40 | 0.8 |
| | ADONA | 4,8-Dioxa-3*H*-perfluorononanoic acid | 919005-14-4 | C7, Short | 0.40 | 0.8 |
| Ether sulfonic acids | PFEESA | Perfluoro(2-ethoxyethane)sulfonic acid | 113507-82-7 | C4, Short | 0.40 | 0.8 |
| | 9Cl-PF3ONS | 9-Chlorohexadecafluoro-3-oxononane-1-sulfonic acid | 756426-58-1 | C8, Long | 0.40 | 0.8 |
| | 11Cl-PF3OUdS | 11-Chloroeicosafluoro-3-oxaundecane-1-sulfonic acid | 763051-92-9 | C10, Long | 0.40 | 0.8 |
| Fluorotelomer carboxylic acids | 3:3FTCA | 3-Perfluoropropyl propanoic acid | 356-02-5 | C6, Short | 0.50 | 0.4 |
| | 5:3FTCA | 2*H*,2*H*,3*H*,3*H*-Perfluorooctanoic acid | 914637-49-3 | C8, Short | 2.5 | 2 |
| | 7:3FTCA | 3-Perfluoroheptyl propanoic acid | 812-70-4 | C10, Long | 2.5 | 2 |

[1]Table ordered by class and chain length.

[2]Chain length is calculated based on the number of carbon chains within the chemical structure.

The definition of "long-chain" PFAS, as provided by the Organization for Economic Cooperation and Development[1], is perfluoroalkyl carboxylic acids with eight carbons and greater (i.e., with 7 or more perfluorinated carbons) and (2) perfluoroalkane sulfonates with six carbons and greater (i.e., with 6 or more perfluorinated carbons). Additionally, Buck et al. suggested that PFAS, except for PFCAs and PFSAs, with a perfluoroalkyl chain with 7 or more C atoms, are "long chain" PFAS.[2]

[3]LOD, limit of detection based on EPA 821-R-16-006.

**Table S2.** Extracted internal standards and injection internal standards used for PFAS quantification.

| Full name | Abbreviation | Amount Spiked (ng) |
|---|---|---|
| ***Extracted internal standard*** | | |
| Perfluoro-n-[$^{13}C_4$]butanoic acid | $^{13}C_4$-PFBA | 0.8 |
| Perfluoro-n-[$^{13}C_5$]pentanoic acid | $^{13}C_5$-PFPeA | 0.4 |
| Perfluoro-n-[1,2,3,4,6-$^{13}C_5$]hexanoic acid | $^{13}C_5$-PFHxA | 0.2 |
| Perfluoro-n-[1,2,3,4-$^{13}C_4$]heptanoic acid | $^{13}C_4$-PFHpA | 0.2 |
| Perfluoro-n-[$^{13}C_8$]octanoic acid | $^{13}C_8$-PFOA | 0.2 |
| Perfluoro-n-[$^{13}C_9$]nonanoic acid | $^{13}C_9$-PFNA | 0.1 |
| Perfluoro-n-[1,2,3,4,5,6-$^{13}C_6$]decanoic acid | $^{13}C_6$-PFDA | 0.1 |
| Perfluoro-n-[1,2,3,4,5,6,7-$^{13}C_7$]undecanoic acid | $^{13}C_7$-PFUnA | 0.1 |
| Perfluoro-n-[1,2-$^{13}C_2$]dodecanoic acid | $^{13}C_2$-PFDoA | 0.1 |
| Perfluoro-n-[1,2-$^{13}C_2$]tetradecanoic acid | $^{13}C_2$-PFTeDA | 0.1 |
| Perfluoro-1-[2,3,4-$^{13}C_3$]butanesulfonic acid | $^{13}C_3$-PFBS | 0.2 |
| Perfluoro-1-[1,2,3-$^{13}C_3$]hexanesulfonic acid | $^{13}C_3$-PFHxS | 0.2 |
| Perfluoro-1-[$^{13}C_8$]octanesulfonic acid | $^{13}C_8$-PFOS | 0.2 |
| 1*H*,1*H*,2*H*,2*H*-Perfluoro-1-[1,2-$^{13}C_2$]hexane sulfonic acid | $^{13}C_2$-4:2FTS | 0.4 |
| 1*H*,1*H*,2*H*,2*H*-Perfluoro-1-[1,2-$^{13}C_2$]octane sulfonic acid | $^{13}C_2$-4:2FTS | 0.4 |
| 1*H*,1*H*,2*H*,2*H*-Perfluoro-1-[1,2-$^{13}C_2$]decane sulfonic acid | $^{13}C_2$-8:2FTS | 0.4 |
| Perfluoro-1-[$^{13}C_8$]octanesulfonamide | $^{13}C_8$-PFOSA | 0.2 |
| N-methyl-$d_3$-perfluoro-1-octanesulfonamide | $D_3$-NMeFOSA | 0.2 |
| N-ethyl-$d_5$-perfluoro-1-octanesulfonamide | $D_5$-NEtFOSA | 0.2 |
| N-methyl-$d_3$-perfluoro-1-octanesulfonamidoacetic acid | $D_3$-NMeFOSAA | 0.4 |
| N-ethyl-$d_5$-perfluoro-1-octanesulfonamidoacetic acid | $D_5$-NEtFOSAA | 0.4 |
| N-methyl-$d_7$-perfluorooctanesulfonamidoethanol | $D_7$-NMeFOSE | 2.0 |
| N-ethyl-$d_9$-perfluorooctanesulfonamidoethanol | $D_9$-NEtFOSE | 2.0 |
| Tetrafluoro-2-heptafluoropropoxy-13C3-propanoic acid | $^{13}C_3$-HFPO-DA | 0.8 |
| ***Injection internal standards*** | | |
| Perfluoro-n-[2,3,4-$^{13}C_3$]butanoic acid | $^{13}C_3$-PFBA | 0.4 |
| Perfluoro-n-[1,2-$^{13}C_2$]hexanoic acid | $^{13}C_2$-PFHxA | 0.2 |
| Perfluoro-n-[1,2,3,4-$^{13}C_4$]octanoic acid | $^{13}C_4$-PFOA | 0.2 |
| Perfluoro-n-[1,2,3,4,5-$^{13}C_5$] nonanoic acid | $^{13}C_5$-PFNA | 0.1 |
| Perfluoro-n-[1,2-$^{13}C_2$]decanoic acid | $^{13}C_2$-PFDA | 0.1 |
| Perfluoro-1-hexane[$^{18}O_2$]sulfonic acid | $^{18}O_2$-PFHxS | 0.2 |
| Perfluoro-n-[1,2,3,4-$^{13}C_4$]octanesulfonic acid | $^{13}C_4$-PFOS | 0.2 |

**Figure S1.** Sample flow chart.

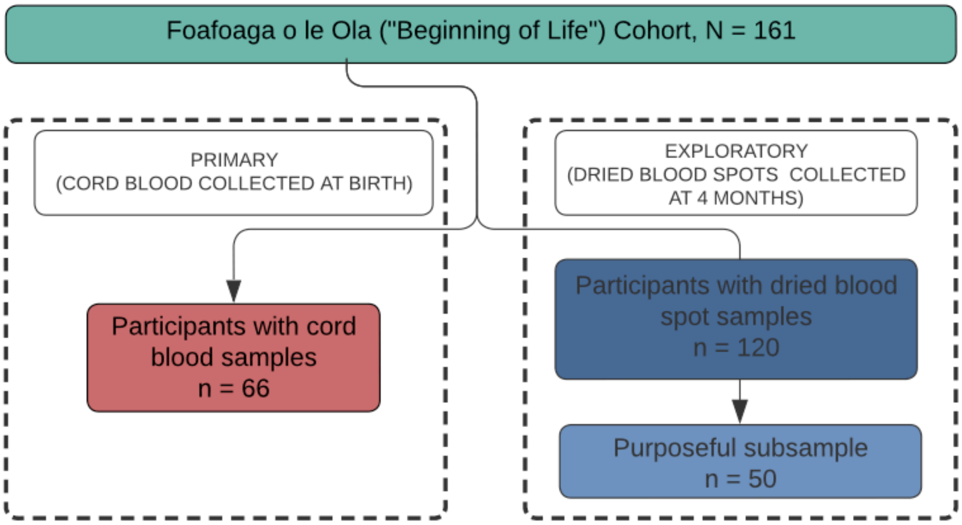

**Table S3.** Expanded sample characteristics with full cohort and subsample comparisons.

| Characteristic | Total sample, N = 161 | Cord blood sample, n = 66 | DBS (4 months) sample, n = 50 |
|---|---|---|---|
| **Maternal characteristics** | | | |
| Maternal age, years | 26.73 (5.71) | 27.33 (5.91) | 27.50 (6.37) |
| *Missing* | 7 | 2 | 0 |
| Maternal BMI, $kg/m^2$ | 34.04 (6.69) | 35.58 (5.80) | 33.87 (5.35) |
| *Missing* | 41 | 7 | 5 |
| Maternal education, years | | | |
| *Less than primary school* | 7 (4.5%) | 2 (3.1%) | 3 (6.0%) |
| *Primary school completed* | 19 (12%) | 8 (12%) | 5 (10%) |
| *Secondary school/college completed* | 90 (57%) | 36 (55%) | 29 (58%) |
| *University completed* | 40 (25%) | 18 (28%) | 13 (26%) |
| *Post-graduate training* | 1 (0.6%) | 1 (1.5%) | 0 (0%) |
| *Missing* | 4 | 1 | 0 |
| Maternal relationship status | | | |
| *Never married* | 15 (9.6%) | 3 (4.6%) | 3 (6.0%) |
| *Currently married* | 98 (63%) | 43 (66%) | 34 (68%) |
| *Cohabitating* | 43 (28%) | 19 (29%) | 13 (26%) |
| *Missing* | 5 | 1 | 0 |
| Census region | | | |
| *AUA* | 77 (49%) | 39 (60%) | 26 (52%) |
| *NWU* | 73 (46%) | 26 (40%) | 22 (44%) |
| *ROU* | 2 (1.3%) | 0 (0%) | 2 (4.0%) |
| *AUA/NWU* | 2 (1.3%) | 0 (0%) | 0 (0%) |
| *AUA/ROU* | 3 (1.9%) | 0 (0%) | 0 (0%) |
| *Missing* | 4 | 1 | 0 |

| Characteristic | Total sample, N = 161 | Cord blood sample, n = 66 | DBS (4 months) sample, n = 50 |
|---|---|---|---|
| **Pregnancy and birth information** | | | |
| Gravidity | 1.97 (1.89) | 1.86 (1.87) | 1.78 (1.62) |
| *Missing* | 12 | 3 | 1 |
| Parity | 1.94 (1.72) | 1.90 (1.59) | 2.07 (1.53) |
| *Missing* | 55 | 25 | 21 |
| Gestational age at birth, weeks | 39.95 (0.83) | 40.08 (0.67) | 39.96 (0.36) |
| *Missing* | 17 | 5 | 3 |
| Birth mode | | | |
| *Vaginal* | 148 (92%) | 59 (89%) | 48 (96%) |
| *Cesarean section* | 13 (8.1%) | 7 (11%) | 2 (4.0%) |
| Infant birth length (cm) | 53.61 (5.90) | 54.73 (5.15) | 52.25 (7.82) |
| *Missing* | 49 | 17 | 14 |
| Infant birth weight (kg) | 3.54 (0.48) | 3.60 (0.48) | 3.47 (0.48) |
| *Missing* | 12 | 2 | 0 |
| Infant sex | | | |
| *Female* | 69 (44%) | 29 (45%) | 27 (54%) |
| *Male* | 88 (56%) | 36 (55%) | 23 (46%) |
| *Missing* | 4 | 1 | 0 |

**Figure S2.** Heat map depicting correlation structure of cord blood PFAS data (n = 66).

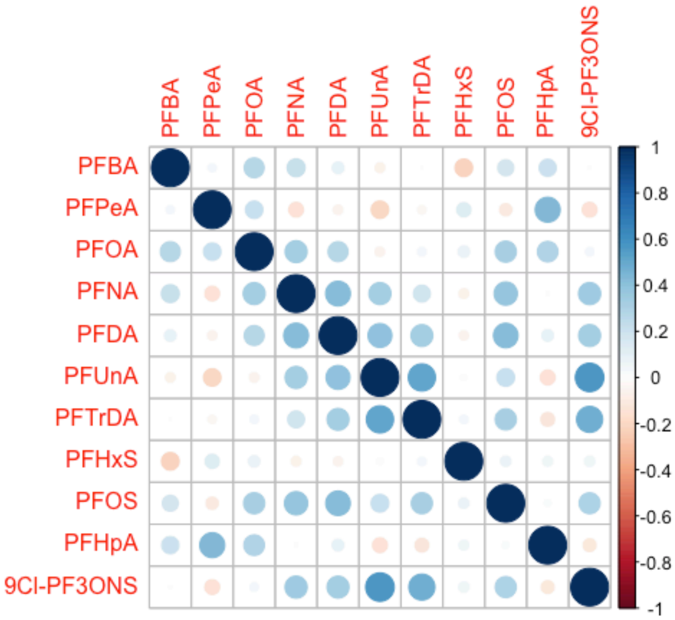


Plot depicts correlation structure (Kendall's Tau [complete data] or Kendall's Tau-B [censored data]) between PFAS levels detected in cord blood. Darker color intensity and larger circle size indicate stronger correlation. Off-diagonal correlations range from -0.18 to 0.57.

**Figure S3.** Heatmap depicting correlation structure of dried blood spot (4 months) PFAS data (n = 50).

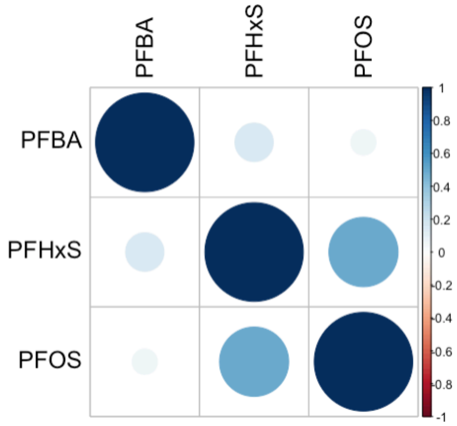


Plot depicts correlation structure (Kendall's Tau-B) between PFAS levels detected in dried blood spots. Off-diagonal correlations range from 0.06 to 0.49.

**Table S4.** Results of regression using maximum likelihood estimation (MLE) examining associations between natural log-transformed cord blood PFAS levels (dependent variable) and census region (NWU vs. AUA) while controlling for infant age, sex, *and socioeconomic status* (n=39).

| Analyte | $\hat{\beta}$ | 2.5th | 97.5th | $exp(\hat{\beta})$ | % Change | p |
|---|---|---|---|---|---|---|
| PFBA | -0.242 | -0.568 | 0.084 | 0.785 | -21.494 | 0.146 |
| PFHxS | -0.236 | -0.560 | 0.088 | 0.790 | -21.022 | 0.153 |
| PFOS | -0.258 | -0.582 | 0.066 | 0.773 | -22.740 | 0.118 |

$\hat{\beta}$ = regression estimate for natural log transformed dependent variable (PFAS analyte).

$exp(\hat{\beta})$ = exponentiated regression estimate for natural log transformed dependent variable (PFAS analyte); in NWU infants compared to AUA infants, the value of y (PFAS analyte) is $exp(\hat{\beta})$ times the original value while holding infant age, sex, and number of socioeconomic resources constant.

% Change = percent increase or decrease in y (PFAS analyte level) in NWU infants compared to AUA infants while holding infant age, sex, and number of socioeconomic resources constant; calculated as % Change = 100 * (exp(Est.)-1).

$2.5^{th}$ = lower bound of 95% confidence interval; $97.5^{th}$ = upper bound of 95% confidence interval.

**Figure S4.** Sina with violin plots describing dried blood spot (4 months) PFAS levels by breastfeeding status at 4 months post-birth (n = 47).

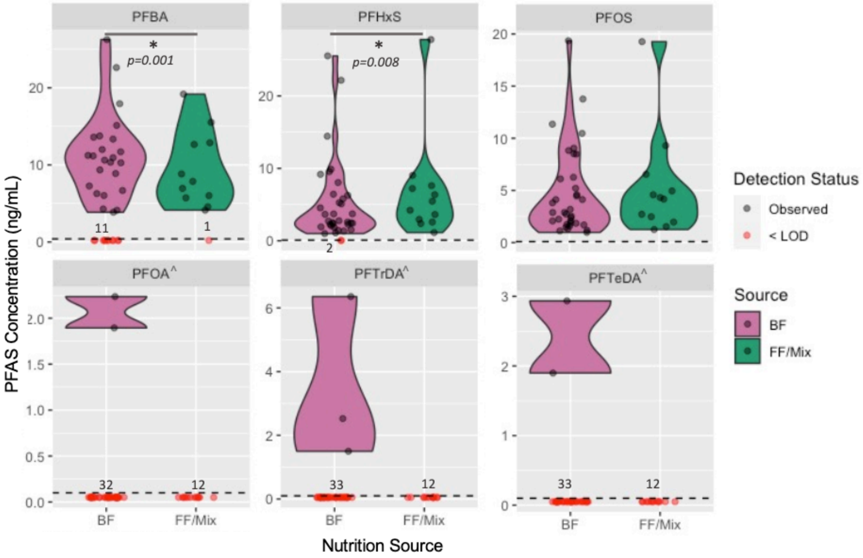


BF = exclusively breastfed at 4 months post-birth, n = 35; FF/Mix = formula- or mixed-fed at 4 months post-birth, n = 12. Formula and mixed-fed nutrition source combined due to sample size. Dashed line represents analyte-specific LOD; black dots indicate observed values; red dots indicate < LOD values (depicted graphically as LOD/2); numbers overlayed on plot facets indicate the number of group-specific < LOD values; violins capture only observed data density. Symbol * indicates statistically significant differences by nutrition source; symbol ^ indicates analytes that were not tested due to high percentage of < LOD values. Figure corresponds with results from Table S4a.

**Table S5.** Results of regression using maximum likelihood estimation (MLE) examining associations between natural log-transformed cord blood PFAS levels (dependent variable) and breastfeeding status (formula-fed or mixed-fed vs. exclusive breastfeeding as reference) while controlling for infant age and sex (Table S4a) and infant age, sex, and socioeconomic resources (Table S4b).

**Table S4a.** Model adjusts for infant age and sex (n=47).

| **Analyte** | $\hat{\beta}$ | **2.5th** | **97.5th** | $exp(\hat{\beta})$ | **% Change** | **p** |
|---|---|---|---|---|---|---|
| PFBA | 0.601 | 0.253 | 0.949 | 1.825 | 82.451 | **<u>0.001</u>** |
| PFHxS | 0.468 | 0.121 | 0.815 | 1.597 | 59.688 | **<u>0.008</u>** |
| PFOS | 0.147 | -0.201 | 0.494 | 1.158 | 15.778 | 0.408 |

**Table S4b.** Model adjusts for infant age, sex, *and number of socioeconomic resources* (n=38).

| **Analyte** | $\hat{\beta}$ | **2.5th** | **97.5th** | $exp(\hat{\beta})$ | **% Change** | **p** |
|---|---|---|---|---|---|---|
| PFBA | 0.723 | 0.330 | 1.117 | 2.061 | 106.061 | **<u>0.0003</u>** |
| PFHxS | 0.627 | 0.235 | 1.019 | 1.872 | 87.199 | **<u>0.002</u>** |
| PFOS | 0.295 | -0.097 | 0.687 | 1.343 | 34.313 | 0.140 |

Note, Table S4a is identical to Table 4; retained here to facilitate comparison across the two analyses.
Formula and mixed-fed nutrition source combined due to sample size.
$\hat{\beta}$ = regression estimate for natural log transformed dependent variable (PFAS analyte).
$exp(\hat{\beta})$ = exponentiated regression estimate for natural log transformed dependent variable (PFAS analyte); in formula or mixed fed infants compared to breastfed infants, the value of y (PFAS analyte) is $exp(\hat{\beta})$ times the original value while holding covariates constant.
% Change = percent increase or decrease in y (PFAS analyte level) in formula or mixed fed infants compared to breastfed infants while holding covariates constant; calculated as % Change = 100 * (exp(Est.)-1).
$2.5^{th}$ = lower bound of 95% confidence interval; $97.5^{th}$ = upper bound of 95% confidence interval.
Bolded p-values indicate statistically suggestive associations (p<0.05) while bolded and underlined p-values indicate statistically significant associations (p<0.025).
Table S4a corresponds with Figure S3.

### Abbreviations

PFAS, Per- and polyfluoroalkyl substances
DBS, Dried blood spot
LOD, Limit of detection
ng, Nanograms
mL, Milliliter
BF, Exclusively breastfed
FF/Mix, Formula fed or mixed fed
